# The Incidence and Severity of COVID-19 in Adult Professional Soccer Players

**DOI:** 10.1101/2020.10.27.20220400

**Authors:** Eduard Bezuglov, Artemii Lazarev, Evgeniy Achkasov, Vladimir Khaitin, Larisa Romanova, Mikhail Butovskiy, Vladimir Khokhlov, Maxim Tsyplenko, Alexander Linskiy, Petr Chetverikov, Magomedtagir Sugaipov, Arseniy Petrov, Oleg Talibov

**Affiliations:** Department of Sport Medicine and Medical Rehabilitation, Sechenov First Moscow State Medical University (Sechenov University), Moscow, Russia; (E. Achkasov); Federal Research and Clinical Center of Sports Medicine and Rehabilitation of Federal Medical Biological Agency, Moscow, Russia; High Performance Sport Laboratory, Moscow Witte University, Moscow, Russia; (E. Bezuglov); A.I. Burnazyan Federal Medical and Biophysical Center, Moscow, Russia; FC Zenit, St. Petersburg, Russia; (V. Khaitin); Head center for hygiene and epidemiology of the Federal Medical Biological Agency, Moscow, Russian Federation; Academy of Postgraduate Education under the Federal State Budgetary Unit “Federal Scientific and Clinical Center for Specialized Medical Assistance and Medical Technologies of the Federal Medical Biological Agency”, Moscow, Russian Federation; (L. Romanova); FC Rubin, Kazan, Russia; (M. Butovskiy); FC Rostov, Rostov-on-Don, Russia; (V. Khokhlov); FC Tambov, Tambov, Russia; (M. Tsyplenko); FC Sochi, Sochi, Russia; (A. Linskiy); FC Orenburg, Orenburg, Russia; (P. Chetverikov); FC Akhmat, Grozniy, Russia; (M. Sugaipov); University of Göttingen, Göttingen, Germany; (A. Petrov); Moscow State University of Medicine and Dentistry, Moscow, Russia; (O. Talibov)

**Keywords:** COVID-19, elite professional soccer, infectious disease

## Abstract

**Introduction:** At present, there are no data regarding the incidence and clinical course of COVID-19 among professional soccer players, and the studies examining putative complications of COVID-19 infections are probabilistic. Thus, examining the incidence of COVID-19 and various aspects of its clinical course in a group of adult professional soccer players would be of great practical interest.

**Methods:** The incidence, clinical course, and severity of COVID-19 infection, as well as the duration of treatment and return to play were studied by the questioning of the team physicians and medical records assessment in the group of adult professional soccer players representing the clubs of the Russian Premier-League (RPL) during the period of championship resumption from 01.04.2020 until 20.09.2020.

**Results:** COVID-19 infection was detected in 103 soccer players in the course of COVID-19 screening. This number comprises 14.5% of all soccer players which were on the rosters of RPL soccer teams and which were subject to regular COVID-19 testing.

The asymptomatic course was observed in 43.7% of cases (n=45). These players were isolated and their clinical condition was monitored closely. Clinical symptoms were observed in 56.3% of cases (n=58), the most common symptoms being fatigue, headache, fever, and anosmia.

**Conclusions:** COVID-19 infection was commonly diagnosed among adult professional soccer players continuously residing in Russia. However, the majority of infections had a mild course and did not impair return to regular exercise.

## Introduction

In December 2019 the Chinese authorities declared the appearance of a novel Coronavirus in the city of Wuhan, which causes a severe respiratory infection [1]. In February 2020 the International Commission on Taxonomy of Viruses officially named the virus causing the current outbreak of the coronavirus disease “SARS-CoV-2”, and the disease caused by this virus has been designated as “COVID-19” [2].

On February 28^th^ the WHO raised COVID-19 threat assessment to its highest level. The COVID-19 outbreak became a major challenge for world health. Around 30 million people got infected with COVID-19 since the beginning of this year. More than 900.000 deceased [3].

Various therapeutic approaches for COVID-19 treatment are utilized, although their efficacy and safety are dubious. The main hope to counter the spread of infection would be a vaccine. Currently, more than 169 candidate vaccines are in development across the world, 26 candidates have reached trials in humans [4]. Nonetheless, widespread safe and efficacious vaccines against COVID-19 are not expected until mid-2021.

The mortality and frequency of complicated disease course due to COVID-19 infection increase with age. The virus most heavily affects elderly patients with comorbidities. The mortality varies between 1.3-8% in the 50-80 years age group, and reaches approximately 15% in the age group above 80 years [2].

Mortality in the age group 18-35 years is significantly lower and comprises approximately 0.2%.

COVID-19 complications such as pulmonary fibrosis, cardiac and hepatic consequences are actively studied [5].

Until now, the course of COVID-19 infection and its impact on athletic performance has not been studied in professional athletes.

Nearly all soccer events were cancelled since March 2020 due to the quick spreading of the global COVID-19 pandemic. The majority of soccer players had to cease training.[6] [7]. During May-June 2020 sports events were resumed in several countries.

However, the events took place without fans in the majority of cases. In all cases when the events were resumed, the organizing sports Leagues developed strict prevention and control measures to minimize the risk of infection for the participants.

The key elements of these measures are as follows: close monitoring of the infection rates by PCR-tests; wearing of individual face masks and gloves; surface and skin disinfection; adherence to social distancing guidelines.

Nevertheless, media reports covering new infections among soccer players illustrate that as efficient as they are, these prevention measures cannot completely rule out the possibility of COVID-19 spreading in such a numerous population as soccer players. However, there is presently no data regarding the incidence and clinical course among professional soccer players, and the studies examining putative complications of COVID- 19 infections are probabilistic.

Thus, examining various aspects of COVID-19 incidence and clinical course in a group of adult professional soccer players would be of great practical importance.

## Materials and methods

The incidence, clinical course, and severity of COVID-19 infection, as well as the duration of treatment and recovery before return to play, were studied by the questioning of the team physicians and medical records assessment in the group of adult professional soccer players representing the clubs of the Russian Premier-League (RPL) during the period of championship resumption from 01.04.2020 until 20.09.2020.

The data of 710 soccer players who were on the rosters of 16 RPL clubs and 2 National Football League teams (second-tier soccer league in Russia) were included in the analysis.

According to the RPL COVID-19 regulation, each player registered for a soccer match has to undergo COVID-19 screening by submitting a throat swab for a PCR test 3 days before the first play, and once weekly thereafter [8].

Without this test, the special QR-code to access the event would not be issued.

Testing is only performed in laboratories certified by the Federal Service for surveillance on consumer rights protection “Rospotrebnadzor” - which is a central Russian governmental entity responsible for sanitary and epidemiological surveillance.

All positive test results are automatically submitted to a centralized database and then being revised in special reference-laboratories.

Test results were available within 24-48 hours in the majority of cases.

According to the Russian quarantine rules, all individuals tested positive for COVID-19 have to be isolated for 14 days regardless of their clinical symptoms. The quarantine can be lifted only after receiving 2 negative PCR-test results performed within at least 24 hours.

Medical records of athletes diagnosed with COVID-19 by a PCR test were studied. Disease course (symptomatic/asymptomatic), the frequency of pulmonary involvement, and the severity of pulmonary lesions were assessed. The prevalence of distinct clinical findings, the therapeutic approaches, the duration of treatment, and recovery before return to play were included in the analysis.

The database was created in Microsoft Excel software, statistical analysis has been performed utilizing the IBM SPSS 23.0 (Armonk, USA). Kolmogorov-Smirnov test was performed to determine whether the data were distributed normally. Descriptive statistics and frequency analysis were used to characterize the sample (mean, SD, min., max.). Mann-Whitney U-test was performed to compare the duration of treatment and recovery before return to play between athletes with and without pulmonary lesions. Results were considered statistically significant at p < 0.05.

The study was approved by the local ethics committee. Players provided written informed consent to participate in the study. It was explained to them that the use of their medical documentation only served for scientific purposes.

## Results

COVID-19 infection was detected in 103 soccer players in the course of COVID-19 screening (average age– 25,1 ± 4,3 years, height – 183,7 ± 6,3 cm, weight – 76,6 ± 7,0 kg, BMI – 22,7 ± 1,4).

This number comprises 14.5% of all soccer players which were on the rosters of RPL soccer teams and which were subject to regular COVID-19 testing.

Halfbacks were most frequently infected – 40.8% of all (n=42), goalkeepers were the least frequently infected players – 11.7% of all (n=12). 29.1% (n=30) of the infected players were full-backs, 18.4% (n=19) were forwards.

The asymptomatic course was observed in 43.7% of cases (n=45). These players were isolated and their clinical condition was monitored closely.

Clinical symptoms were observed in 56.3% of cases (n=58), the most common symptoms being fatigue, headache, fever, and anosmia (Table 1).

**Table 1.**
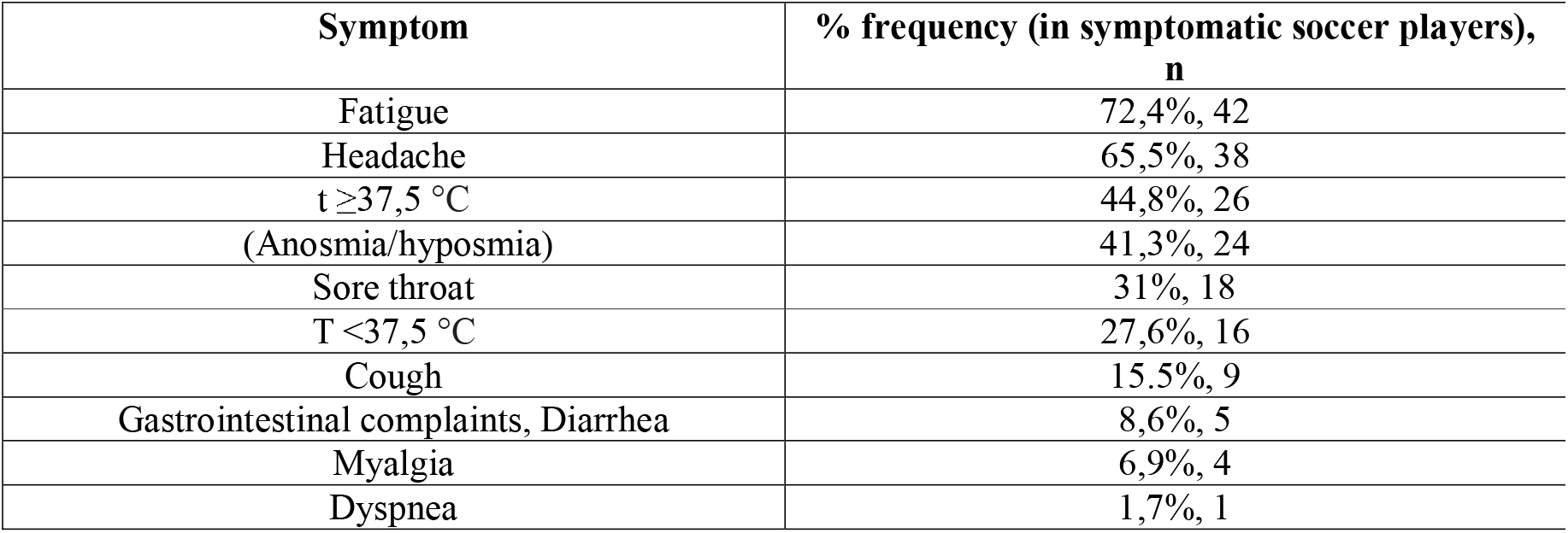
The most frequent clinical symptoms in soccer players with COVID-19 infection.

| Symptom | % frequency (in symptomatic soccer players),<br>n |
| --- | --- |
| Fatigue | 72,4%, 42 |
| Headache | 65,5%, 38 |
| $t \geq 37,5$ °C | 44,8%, 26 |
| (Anosmia/hyposmia) | 41,3%, 24 |
| Sore throat | 31%, 18 |
| $T < 37,5$ °C | 27,6%, 16 |
| Cough | 15,5%, 9 |
| Gastrointestinal complaints, Diarrhea | 8,6%, 5 |
| Myalgia | 6,9%, 4 |
| Dyspnea | 1,7%, 1 |

CT scans have shown pulmonary lesions in 36,2% (n=21) of symptomatic soccer players. Pulmonary lesions were detected in 23.3% (n=24) of players with positive test results (in 3 cases, pulmonary lesions were detected in asymptomatic players).

Pulmonary lesion size was derived from CT images. Less than 10% of lung parenchyma were involved in 70.9% (n=17) of cases, 11-20% in 16.6% (n=4), and 20-29% in 8.3% (n=2) of cases. Lesions involving more than 30% of lung parenchyma were detected only once (4.2%; n=1).

Pulmonary lesions were most commonly associated with the following symptoms: fatigue (76.2%, n=16), headache (71.4%, n=15), t ≥ 37.5 °C (52,4%, n=11).

Medical records of 59.2% of players (n=61) were available for assessment. Players were clustered by the drug received (Table 2).

**Table 2.** Drugs utilized for the treatment of COVID-19 infection.

| Drugs | Prevalence (% , n) |
| --- | --- |
| Antivirals | 85,2%, 52 |
| Antibiotics | 83,6%, 51 |
| Vitamins | 59%, 36 |
| Anticoagulants | 26,2%, 16 |
| Interferons | 14,8%, 9 |
| Anti-malaria agents | 11,5%, 7 |
| NSAIDs | 6,6%, 4 |

The average treatment duration of symptomatic soccer players without pulmonary manifestations of the disease was 14,4 ± 4,8 days, and 17,4 ± 3,5 days in those with pulmonary lesions.

Pulmonary lesions were not identified as a risk factor contributing to a longer duration of treatment (logistic regression, p = 0.09) or delaying the resumption of training as a regular player in the respective soccer team (logistic regression, p = 0.17).

Significant difference in treatment duration of players with and without pulmonary lesions was registered (p = 0.022, Mann-Whitney), and no significant difference has been observed in duration of recovery before return to play (p = 0.29, Mann-Whitney). Treatment duration was significantly longer in the players with pulmonary lesions.

No complications which required pulmonary ventilation have been registered in any of the cases.

All players underwent a thorough medical examination, which is obligatory for all professional athletes in Russia, before returning to training. No cardiovascular, pulmonary, and hematological abnormalities were detected in players who recovered from COVID-19 infection.

The average duration of recovery before return to play was 19,0 ± 5,3days. The recovery period lasted 20,7 ± 7,3 days (non-normal distribution) in those who suffered COVID- pneumonia and 18,4 ± 4,4 days (non-normal distribution) in those who had no pulmonary manifestations. No significant correlation between COVID-pneumonia and the duration of recovery period was detected (Mann-Whitney, p = 0,32).

The disease severity was assessed by the number of clinical findings. Distribution of age and clinical findings is non-normal; thus, Spearman’s rank correlation coefficient was utilized. No significant and strong correlation between the age and clinical findings has been detected (p = 0.1; R = 0.18).

Almost all soccer players who had symptomatic COVID-19 disease have already participated in soccer events at the moment of study recruitment. Only two players are currently not active due to expired contracts with their clubs.

## Discussion

We have demonstrated the predominance of asymptomatic forms of the COVID-19 infection among professional soccer players continuously residing in Russia. If symptoms occurred, the course of the disease has always been mild. In all cases, the COVID-19 infection did not lead to the development of severe complications and did not impair return to training after convalescence.

This is the first study of the kind, examining COVID-19 in a population of elite professional soccer players.

It would be interesting to compare the prevalence of symptoms in our study with the data previously described in the literature. Grant et al. have performed a systematic review and a meta-analysis of 148 scientific papers, including a total of 24410 adult patients with COVID-19 disease (age >16 years, average age 49 years) from 9 countries. The most common symptoms described were as follows: fever – 78%, cough – 57%, fatigue – 31%. Headache was observed in 13% of cases, hyposmia – 25%, and myalgia - 17%. The most common symptoms in our study were as follows: fatigue (72.4%), headache (65.5%) and fever > 37.5 degrees centigrade (44,8%). Anosmia/hyposmia was observed in 41.3% of cases. Only 56.3% of soccer players had symptomatic disease, 43.7% were asymptomatic. In contrast to our study, Grant et al. assessed all symptoms in patients with a positive test result. Thus, the symptomatic course of the disease was less commonly observed in our study, which is most probably due to the younger age and excellent health of the soccer players [9].

It would be interesting to compare our results with the findings of the previous studies. Bernheim et al. performed a retrospective study of 121 symptomatic COVID-19 patients from China (average age 45 +- 15.6 years). Pulmonary lesions were observed in 78% of patients. In our study, only 20.4% of soccer players had pulmonary manifestations, which is probably due to timely diagnosis and early treatment start [10].

Meyer et al. demonstrated a relatively low incidence of SARS-CoV-2 infections among German soccer players. During the study period from May to July 2020 (9 weeks), 8 players were tested positive in the first PCR-testing round before the resumption of training sessions, 2 players were tested positive in the third round of PCR-testing, and 22 players remained seropositive throughout the whole season [11].

The numbers reported in this study are much lower than ours. The differences might be explained by several factors.

When club training and competitive matches in Russia were resumed, the fans were allowed to enter the stadium, which facilitates the spread of infection. In contrast to Russia, viewers were not allowed at Bundesliga games [11].

Another factor influencing the frequency of COVID-19 infections among Russian soccer players could be the frequent flying, and thus a large number of close contacts. Each team had at least 20 flights, lasting >2 hours during the study period. Although the teams used charter flights, the risk of infection was nevertheless high.

However, a higher detection rate in Russian players could be due to meticulous PCR-testing done by the physicians’ staff once in a week, which allowed them to detect infections early and take actions to promptly isolate and treat the players who were tested positive.

An important aspect in the treatment of professional athletes is strict adherence to anti-doping regulations. If prohibited substance use is necessary for treatment, an athlete has to apply for a Therapeutic Exemption (TUE) by the respective national anti-doping association. Considering that a uniform COVID-19 treatment method has not been established yet, in some cases it could be problematic to retroactively receive a TUE from the WADA, e.g. after being treated with dexamethasone[12].

However, the treatment-protocols applied to treat soccer players did not require the use of forbidden substances such as dexamethasone. Cardio-toxic drugs such as hydroxychloroquine were not applied as well.

The duration of the recovery period to enable the safe resumption of training is of critical importance for soccer players. It has been assumed, that COVID-19 itself as well as various therapeutics to treat the infection might cause negative effects on organ systems, primarily on the cardio-respiratory system [13]. However, the side effects might be sub-clinical and not appear instantly [14]. The importance to monitor the health of an athlete recovered from COVID-19 has been stressed in previous studies [15].

According to the Russian sports regulations, all professional athletes must undergo a compulsory thorough medical examination twice in a year. It includes a cardiac check-up (ECG, Echo-CG, a cardiac stress-test); respiratory check-up (spirometry, chest radiograph), general and chemistry panel blood tests [16]. These tests are done in specially licensed clinics by the sports medicine physicians. After doing the tests an athlete receives a medical certificate which is to submit to a respective sports federation. Without such a certificate, athletes are not permitted to training or competitions.

All soccer players who recovered from COVID-19 (symptomatic or asymptomatic form) underwent a thorough medical examination. None of them was diagnosed with any pulmonary or cardiac pathologies or exercise intolerance. None was recommended to restrain from physical activity irrespective of its intensity.

Thus, asymptomatic and symptomatic soccer players with mild pulmonary manifestations of the disease did not demonstrate any impairment in respiratory and cardiovascular function or exercise intolerance in the short-term after recovery from COVID-19 infection. The long-term effects of COVID-19 should be the focus of future research.

## Conclusion

COVID-19 infection was commonly diagnosed among adult professional soccer players continuously residing in Russia. However, the majority of infections had a mild course and did not impair return to regular exercise.

## Data Availability

No data are available. All original data obtained from the players are fully confidential.

## What are the findings?

- COVID-19 was often diagnosed among professional soccer players continuously residing in Russia. The percentage of players who tested positive comprised 14.5%.
- Infection with COVID-19 was difficult to evade despite strict compliance with all preventive measures
- The majority of players were asymptomatic or had a mild disease course

## How might it impact on clinical practice in the future?

- The treatment of COVID-19 in professional soccer players usually do not require the usage of drugs from the WADA List of prohibited substances, as well as cardio-toxic drugs.
- Symptomatic soccer players with pulmonary manifestations of the disease do not demonstrate any impairment in respiratory and cardiovascular function or exercise intolerance after recovery from COVID-19 infection. Any long-term effects are not known yet.

